# A discrete choice experiment to understand depression intervention treatment preferences of Kenyan pregnant adolescents

**DOI:** 10.1101/2022.08.07.22278515

**Authors:** Manasi Kumar, Albert Tele, Joseph Kathono, Vincent Nyongesa, Obadia Yator, Shillah Mwaniga, Keng Yen Huang, Mary McKay, Joanna Lai, Marcy Levy, Pim Cuijpers, Matthew Quaife, Jurgen Unutzer

## Abstract

**Background:** Understanding mental health treatment preferences of adolescents and youth is particularly important for interventions to be acceptable and successful. Person-centered care mandates empowering individuals to take charge of their own health rather than being passive recipients of services.

**Methods:** We conducted a discrete choice experiment to quantitatively measure adolescent treatment preferences for different care characteristics and explore tradeoffs between these. A total of 153 pregnant adolescents were recruited from two primary healthcare facilities in the informal urban settlement of Nairobi. We selected eight attributes of depression treatment option models drawn from literature review and previous qualitative work. We created a balanced and orthogonal design to identify main term effects. A total of ten choice tasks were solicited per respondent. We evaluated mean preferences using mixed logit models to adjust for within subject correlation and account for unobserved heterogeneity.

**Results:** Respondents showed a positive preference that caregivers be provided with information sheets, as opposed to co-participation with caregivers. With regards to treatment options, the respondents showed a positive preference for 8 sessions as compared to 4 sessions. With regards to intervention delivery agents, the respondents had a positive preference for facility nurses as compared to community health volunteers. In terms of support, the respondents showed positive preference for parenting skills as compared to peer support. Our respondents expressed negative preferences of ANC service combined with older mothers as compared to adolescent friendly services and of being offered refreshments alone. A positive preference was revealed for combined refreshments and travel allowance over travel allowance or refreshments alone.

**Conclusion:** This study highlights unique needs of this population. Pregnant adolescents value depression care services offered by nurses Participants shared a preference for longer psychotherapy sessions and their preference was to have adolescent centered maternal mental health and child health services within primary care.

## Introduction

The prevalence of depression is high among pregnant women, with worldwide estimates of 11-18% [1,2] and between 15-28% in Lower-and-Middle-Income Countries (LMICs) (4-8). Adolescent mothers usually experience higher rates of prenatal depression as compared adult mothers [5]. Maternal depression negatively impacts the maternal and child health[3,4,6]. In Kenya, pregnant adolescents report mental health problems, difficulty in accessing financial, moral and material support from parents or partners, and stigmatization by health workers when seeking health care[7]. Discrete choice experiments (DCE) enable us to estimate relative preference weights and their corresponding trade-offs to measure what is important to people when choosing to engage in care[8], and though this approach has been tested within mental health field[9], it can more actively be used in low resource contexts to prioritize patient centered care[10]

DCEs offer rigorous and systematic approaches for eliciting preferences for service or product attributes from customers and stakeholder[11]. DCEs allow for estimation of the relative importance of aspects of the service by analyzing trade-offs between attributes made by stakeholders. This method is increasingly applied to healthcare settings to enable patient input for patient-centered care [12] and has been successfully applied for patient preference elicitation in multiple areas of healthcare, including provider-interactions, health service delivery content and format, and treatment options [12].

Patients’ preferences are particularly salient in depression treatment, because multiple efficacious treatments (for example, combination of antidepressants and psychotherapies) and modalities (for example, group and individual) as well as different types of psychotherapies (cognitive-behavioral or relational like IPT) exist. Incorporating individual patients’ preferences into treatment decisions could lead to improved adherence to treatments for depressive disorders in this highly vulnerable group.

The objective of conducting this DCE was to quantitatively measure adolescent depression treatment preferences for different care characteristics and explore tradeoffs between these.

## Methods

### Design

A DCE is a survey design that asks respondents for their utilities [13]. The method is based on random utility theory [14] and Lancaster’s economic theory of value [15]. It is built on the assumptions that health care interventions, services, or policies can be described by their characteristics (called attributes), and that a person’s valuation depends on the levels of these characteristics [16,17].

DCEs ask people to complete a series of hypothetical choice activities to extract this information. Individuals are asked to choose their favorite option among two or more alternatives (e.g., psychological therapies) with varying intervention characteristics in each choice task. Patient preferences can thus be measured as the extent to which each intervention feature influences an individual’s intervention choice.

This survey was created in accordance with the International Society for Pharmacoeconomics recommendations and the Outcomes Research (ISPOR) Conjoint Analysis Task Force checklist for appropriate research techniques for stated-preference studies[18]. The task force’s experimental design guidelines [19] were used in this investigation (see supplementary table 1).

To investigate depression treatment preferences among pregnant teenagers, we employed a DCE, which consists of four stages: identifying and defining attributes and levels, generating choice sets and constructing questionnaires, collecting survey data, and analyzing and explaining the results [15,20]. Figure 1 depicts the DCE’s development process.

**Figure 1.**
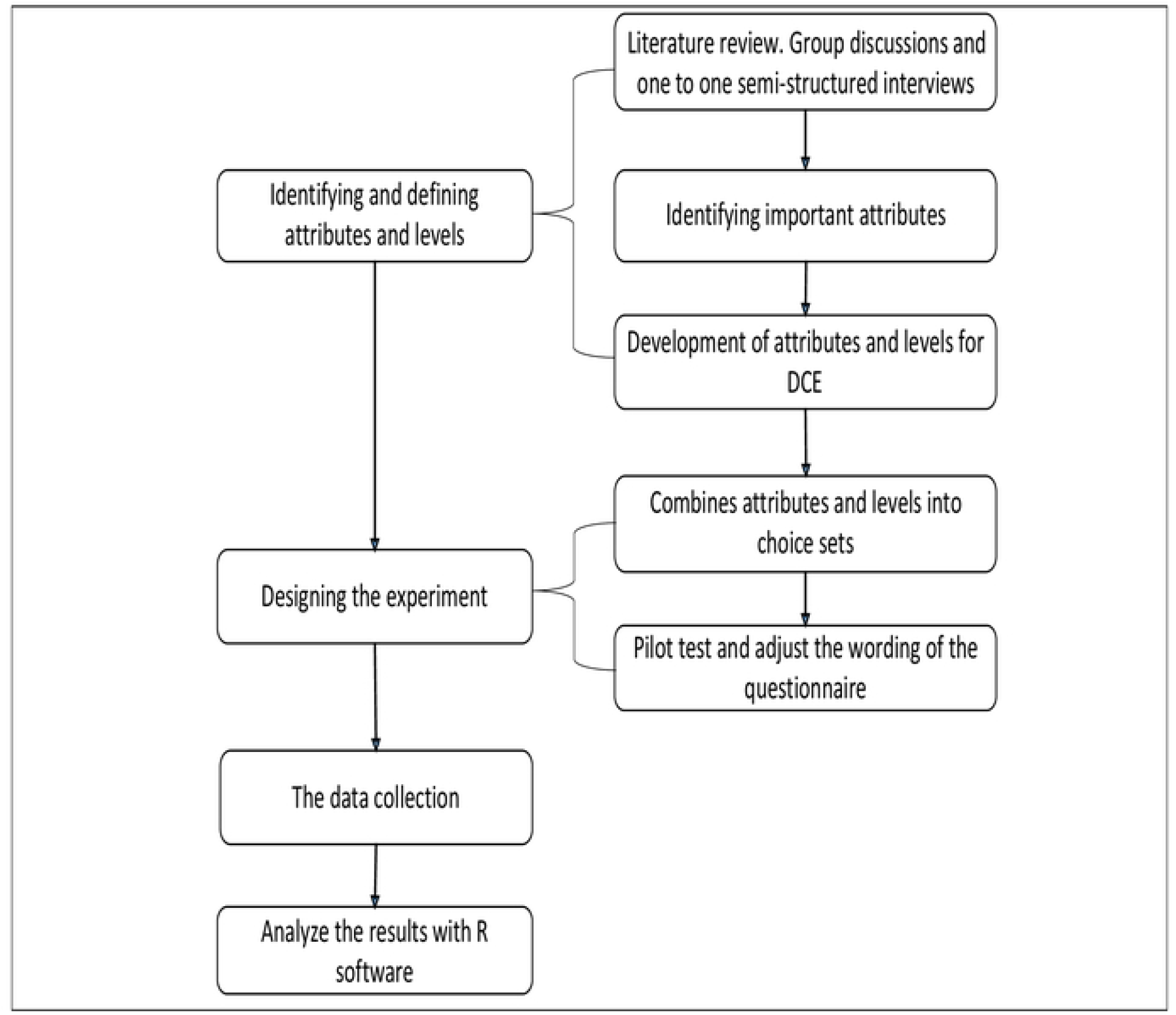
The Development Process of DCE

### Attributes and Levels

A preliminary list of attributes was made by extracting all relevant attributes and levels from health-related DCEs [9,21] and through comprehensive literature review in consultation of with psychologists with experience in adolescent mental health. We supplemented this by conducting semi-structured qualitative interviews with experts in the field of adolescent mental health and health economics (primary health clinicians, nurses and mental health practitioners (n=36), researchers in the field of mental health (n=10), and a health economist) [22,23]. Subsequently, we conducted qualitative semi-structured interviews with 10 purposefully sampled respondents with a history of depression diagnosis. Following a grounded theory approach in the phases of both data collection and analysis[24], we derived lists of actors and factors that may play a part in the search for and selection of depression treatment. The authors reduced the set of potential candidate attributes to a more manageable set of attributes by filtering out double or overlapping attributes. The final list included eight attributes that consisted of; (i) Information delivery, (ii) Participants, (iii) Treatment option, (iv) Intervention delivery, (v) Training, (vi) Support, (vii) Services and (viii) Incentives.

The design was pilot-tested with a selection of the pregnant adolescents who had been participated in the qualitative interviews to refine the survey and to assess the salience of the attributes to the treatment decision. Participants completed DCE questionnaires and participated in a personal cognitive interview as part of the pilot testing. To determine the burden on participants, the number of completed items and the time it took to complete them were recorded. Personal cognitive interviews were utilized to assess participants’ knowledge of the questionnaire’s levels and face validity. The final set of attributes and levels are presented in Table 1.

**Table 1:** Attributes and levels

| Attributes | Level |
| --- | --- |
| 1. Information delivery | Health Facility<br>Individualized |
| 2. Participants | Co-participate with Caregivers<br>Provide information sheets for care-givers |
| 3. Treatment option | 4 sessions for 1.5 hours<br>8 sessions for 1.5 hours |
| 4. Intervention delivery | CHV<br>Facility Nurses |
| 5. Training | Vocational<br>Formal (Back to school) |
| 6. Support | Peer support<br>Parenting skills |
| 7. Services | Adolescent friendly services<br>Combined with older mothers |
| 8. Incentives | Transport KSh. 500<br>Food<br>Both |

We tested multiple-choice elicitation formats and chose to use full-profile tasks between two treatment profiles in which participants indicated which treatment they would prefer to take. This setup allowed for the elicitation of acceptable tradeoffs people were willing to make between different treatment attributes. If the number of attributes is low enough that participants can reasonably complete a full-profile task, this maximizes information about trade-offs [25]. We allowed the participants to select an opt-out option. An example choice task with decision scenario is shown in Figure 2.

**Figure 2.**
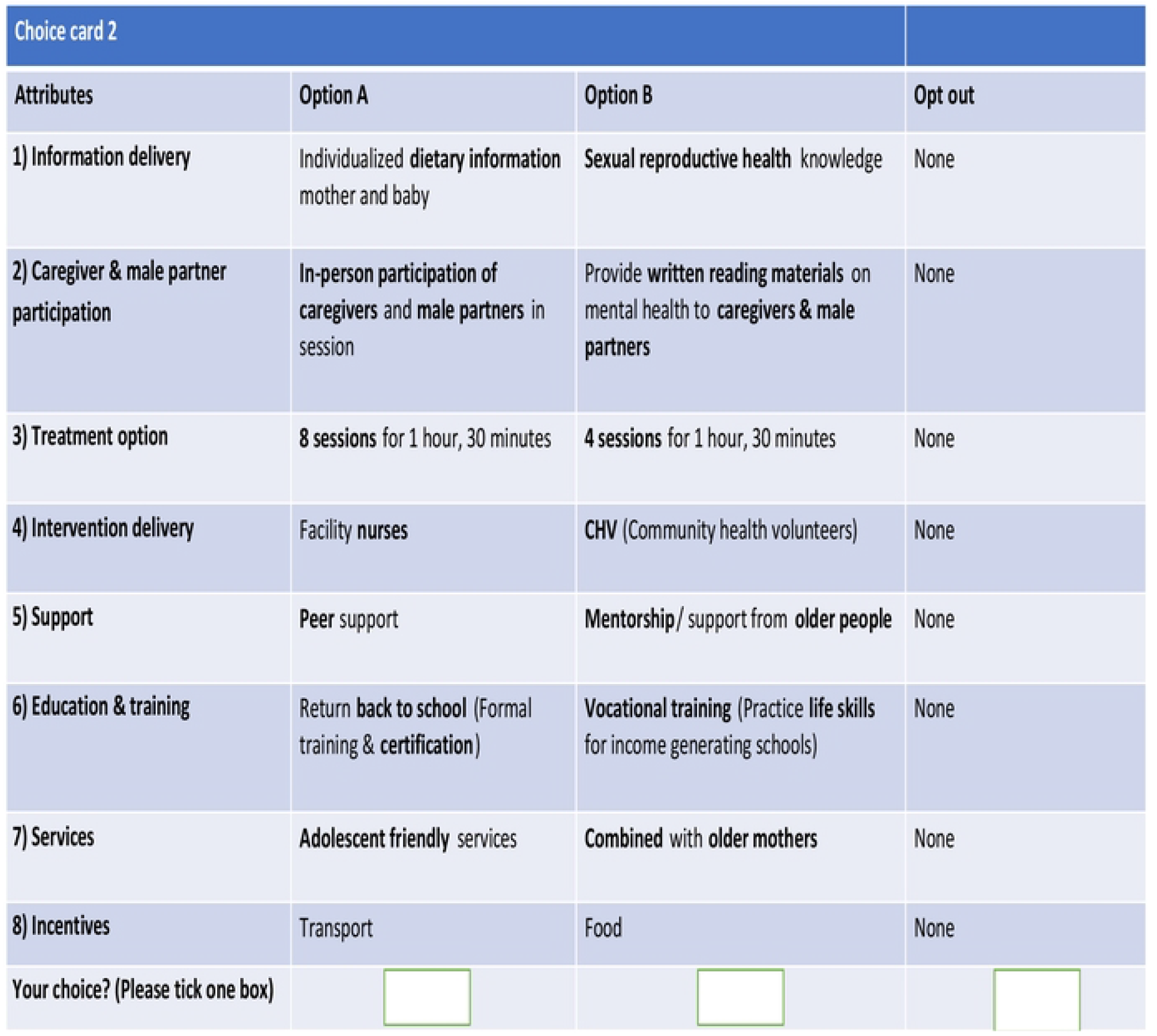
Sample choice card using orthogonal design^*^ *Note: In an orthogonal design all attributes are independent of one another

**Figure 3.**
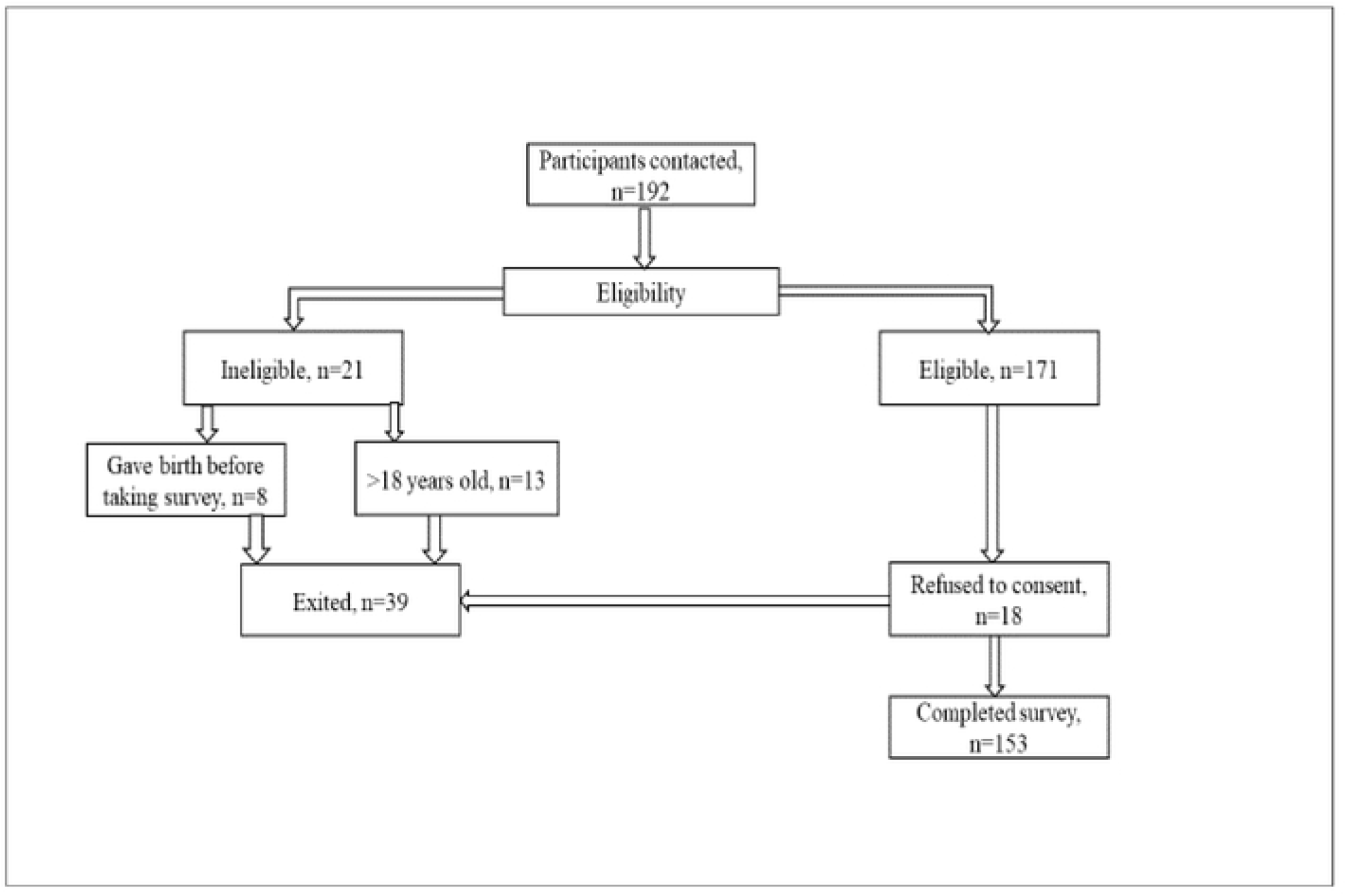
Recruitment Flow Chart

**Figure 4.**
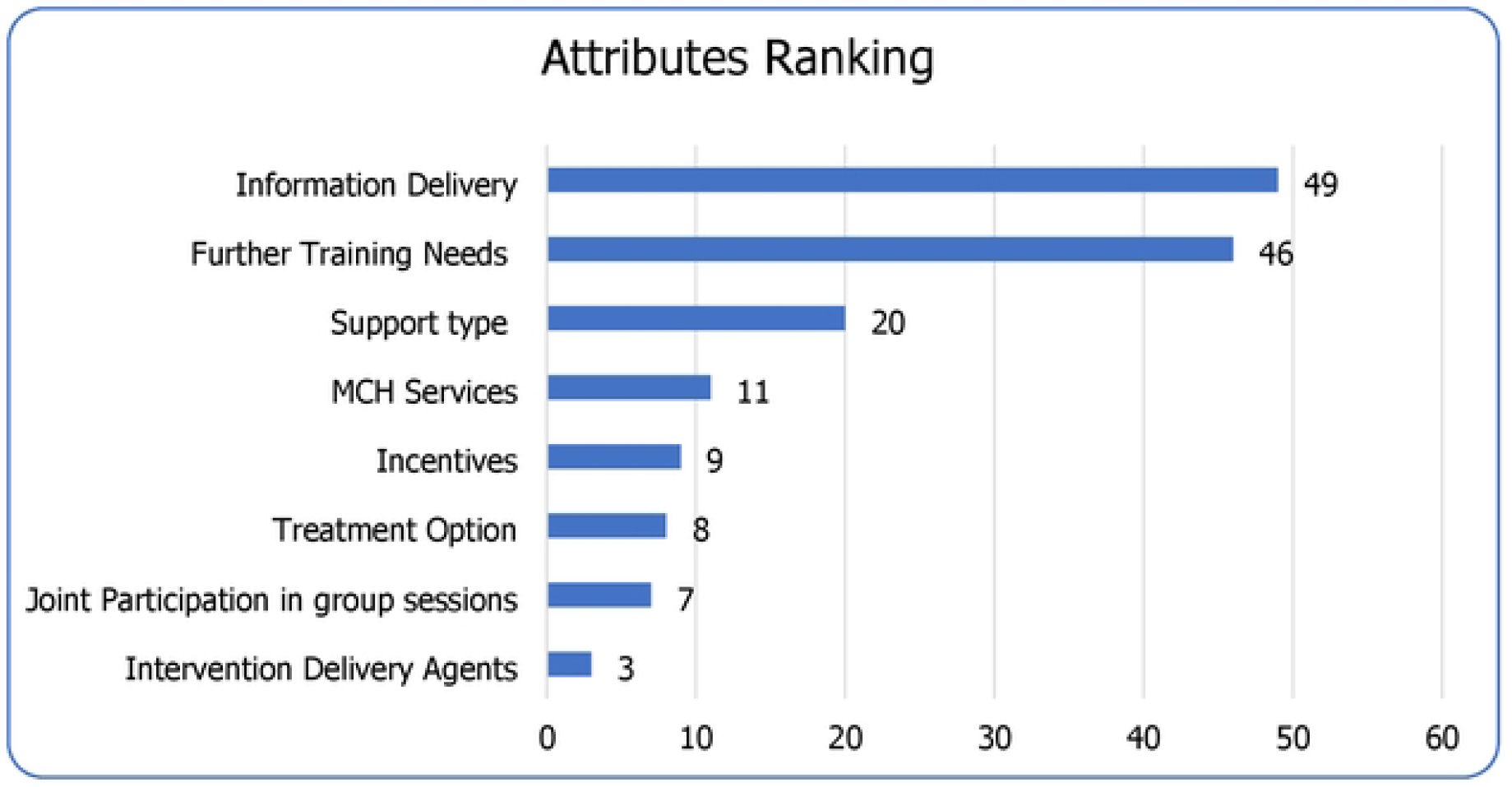
Ranking of attributes

### Experimental Design

We piloted using a fractional factorial design, then analyzed data in a multinomial logistic regression model to generate a Bayesian D-optimal design [26]. D-optimal designs maximize the precision of the estimated parameters given a set number of choice tasks and information on expectations of respondent preferences [27]. We designed ten choice tasks. A repeat task was added to test choice consistency. In addition to discrete choice tasks, participants were asked to directly rank the attributes in order of importance from 1-8.

### Recruitment and Data Collection

Purposive sampling was used to recruit pregnant adolescents aged 14-18 years who were seeking antenatal services at two primary health care centers located in two informal settlements with Nairobi County were recruited from March to June 202. In addition, respondents who had participated in the preliminary study were approached (qualitative interviews and pilot study). The recruitment was carried out by two research assistants and seven CHVs. In Addition, 11 choice scenarios, we administered PHQ-9 to measure self-reported depressive symptoms and also collected information on their socio-demographic profiles. Confirmed pregnancy status, adolescent age 14-18 years, willingness to share their feedback on the DCE, familiarity with Kiswahili and English languages, stable mental health, being in the neighborhood for last one year, willingness to give consent were the exact inclusion criteria.

The sample size estimate in our study is based on Johnson and Orme’s rule of thumb (R Johnson & Orme, 2010; Rich Johnson & Orme, 2003). The calculation formula for the minimal sample size N, according to Johnson and Orme, is provided in the following equation: N ≥ (500 x c)/(a x t) -where N is the number of participants, t is the number of choice tasks (questions), a is the number of alternative scenarios and c is the largest number of attribute levels for any one attribute, and when considering two-way interactions, ‘c’ is equal to the largest product of levels for any two attributes -(500 × 6/ 3 × 8). To account for 10% non-response at least 139 participants is recommended. A tablet-based DCE interview using Dooblo software program [28] was used to capture participant responses.

#### Ethical approval

Ethical approval was obtained from the Kenyatta National Hospital and University of Nairobi Ethics and Research Committee (Approval No. P694/09/2018). All data collection, such as de-identifying data and allowing participants to stop the survey at any time, was done in accordance with ethical standards.

### Statistical Analysis

We analyzed choice data using mixed multinomial logit models [29] to adjust for within-subject correlation [30,31] and account for unobserved preference heterogeneity [29]. The model was estimated using the mlogit command with 500 random Halton draws in R version 4.1.2. No interaction terms were included and we did not perform sub-group analysis. In the mixed logit model, all the attributes were included as effects-coded categorical variables that we assumed to be normally distributed. This assumption was based on convenience because appropriate assumptions for these distributions remain ambiguous [15,25,29]. We chose to use effects coding to account for nonlinearities [25]. The level of each attribute that we expected to be most neutral was used as the omitted or reference attribute parameter. The negative preference (represented by negative coefficient) represents disutility or disliking of that option and positive preference represents liked or beneficial choice/utility (positive coefficient) on mixed logit model.

## Results

### Response rate

A total of 192 participants who were registered in ANC clinics at Nairobi Metropolitan Service’s Kariobangi and Kangemi health centers were contacted and screened for eligibility. Of these, 21 participants did not meet the eligibility criteria, where eight participants gave birth before the study commenced and 13 were aged more than 18 years. Out of 171 eligible participants, 18 refused to consent leaving a total sample size of 153 (Response rate of 89.5%).

### Participant sociodemographic characteristics

Table 2 presents the socio-demographic and other characteristics of the respondents. A total of 153 pregnant adolescent girls participated in the survey The mean age was 17.2 and ranged from 14–18 years. More than three-quarters of participants (79.7%) were single, while the rest were either married or living with a partner. In terms of education, the majority (72.5%) had secondary school level of education. Most respondents (64.1%) were students and the rest were staying at home doing family chores. Participants who were classified as having probable depression (PHQ-9>10) were 43.1% (*95% C*.*I*. 35.3% -51.6%, mean (SD): 8.9 (9.0).

**Table 2:** Socio-demographic Profile of the Respondents

| Variable | Category | Frequency<br>(N=153) | Percentage<br>(%) |
| --- | --- | --- | --- |
| Age | Mean; SD; Range | 17.2; 1.0; 14-18 |  |
| Marital status | Single | 122 | 79.7 |
|  | Married/ co-habiting<br>with a partner | 31 | 20.3 |
| Highest level of education | Primary school | 42 | 27.5 |
|  | Secondary school | 111 | 72.5 |
| Monthly Family Income | <4,999 KES/ under 500<br>USD | 82 | 53.6 |
|  | 5,000-9,999 KES/500-100 USD | 34 | 22.2 |
|  | >10,000 KES/1000<br>USD | 37 | 24.2 |
| Person Living With | Parents | 103 | 67.3 |
|  | Spouse | 32 | 20.9 |
|  | Others | 18 | 11.8 |
| Week of gestation of first ANC visit | < 12 weeks | 25 | 16.3 |
|  | 12-28 weeks | 108 | 70.6 |
|  | >28weeks | 20 | 13.1 |
| Unplanned Pregnancy | Yes | 96 | 62.7 |
|  | No | 57 | 37.3 |
| Attitude of adolescent's partner towards<br>the pregnancy | Positive | 92 | 60.1 |
|  | Negative | 40 | 26.1 |
|  | Ambivalent | 21 | 13.7 |
| Presence of social support | Yes | 120 | 78.4 |
|  | No | 33 | 21.6 |
| Parity | 0 | 14 | 9.2 |
|  | 1 | 113 | 73.9 |
|  | 2+ | 26 | 17.0 |
| Age at Sex Debut | 14-16 Years | 79 | 52.0 |
|  | 17-18 Years | 73 | 48.0 |
|  | *Non-Response* | *1* |  |

### Preference Results

The results from the mixed logit model are presented in Table 3. Given the significant estimates for all but one attribute level information delivery and the alignment of coefficients with a priori expectations, we conclude the DCE was well understood by participants and the modelling method appropriate. The attributes are ranked from the most preferred to the least preferred in terms of the strength of their coefficients.

**Table 3:** Results of mixed multinomial logit model with calculated proportions of positive and negative effects for treatment options

| Variable | Category | $\beta$ | S. D | %<br>Preference | S. E | Sig. |
| --- | --- | --- | --- | --- | --- | --- |
| Information delivery | SRHR information | Ref. |  |  |  |  |
|  | Individualized dietary information | -0.66 | 26.83 | -50.8 | 0.76 | 0.3864 |
| Joint Participation | Co-participate with Caregivers/partners | Ref. |  |  |  |  |
|  | Provide information sheets for care-givers | 6.05 | 16.38 | 64.4 | 0.82 | <0.001 |
| Treatment duration | 4 sessions for 1.5 hours | Ref. |  |  |  |  |
|  | 8 sessions for 1.5 hours | 5.52 | 122.11 | 52.0 | 1.33 | <0.001 |
| Intervention delivery agents | CHV | Ref. |  |  |  |  |
|  | Facility Nurses | 6.04 | 87.78 | 52.8 | 1.27 | <0.001 |
| Further Training Needs | Vocational | Ref. |  |  |  |  |
|  | Formal (Back to school) | -19.29 | 25.77 | -77.3 | 1.13 | <0.001 |
| Support type | Peer support | Ref. |  |  |  |  |
|  | Parenting skills | 19.57 | 113.50 | 56.7 | 1.51 | <0.001 |
| MCH Services | Adolescent friendly services | Ref. |  |  |  |  |
|  | Combined with adult/older women | -37.92 | 107.82 | -63.7 | 1.64 | <0.001 |
| Incentives | Food | -4.69 | 118.83 | -51.6 | 1.74 | 0.0071 |
|  | Transport | 18.39 | 5.53 | 99.9 | 0.81 | <0.001 |
|  | Both | Ref. |  |  |  |  |
| Opt Out |  | -118.10 | 32.58 | -99.9 | -118.10 | <0.001 |

Note: Ref-Reference category; MCH-Maternal and child health care

With regards to services delivery, the respondents had a negative preference of being offered services along with adult mothers at the ANC (63.7%, β=-37.92, *p<0*.*001*) as compared to separate adolescent friendly services. The respondents expressed positive preference for training in parenting skills (56.7%, β=19.6, *p<0*.*001*) as compared to peer-support based skills. In terms of further training needs, there was a negative preference for return to school (77.3%, β=-19.3, *p<0*.*001*) as opposed to livelihood training.

In terms of added incentives to make improve access to IPT sessions, a negative preference for refreshments was expressed (51.6%, β=-4.69, *p<0*.*001*) as compared to provision of combined transport funds and refreshments. Interestingly, our respondents had a positive preference for receiving transport allowance (99.9%, β=18.39, *p<0*.*001*) in comparison of both refreshments and travel allowance.

In terms of joint participation group sessions, the respondents showed a positive preference that the caregivers be provided with information sheets (64.4%, β=6.05, *p<0*.*001*), as opposed to co-participation with caregivers or partners.

When asked who is preferred for running intervention delivery sessions, the respondents had a positive preference for facility nurses (52.8%, β=6.04, *p<0*.*001*) as compared to CHVs.

In terms of intervention delivery, our respondents showed a positive preference of 8 sessions (52.0%, β=5.52, *p<0*.*001*) as compared to 4 sessions.

Participants did not show significant differences in preference for information delivery. Respondents were less likely to opt out of the choices with 99.9% of them opting to choose one of the two options provided.

Table 3: Results of mixed multinomial logit model with calculated proportions of positive and negative effects for treatment options

### Ranking of attributes

Our respondents ranked Information delivery as the first, treatment duration option second, Support type third, Joint Participation fourth, MCH Services fifth, Incentives sixth, and Intervention Delivery Agents seventh and Further training needs as eighth.

## Discussion

This study assessed depression treatment preferences among pregnant adolescent girls in informal urban setting. Consistent with prior expectations, information delivery and treatment options were the most important attributes. Our respondents showed a positive preference that caregivers be provided with information sheets, as opposed to co-participation in therapy sessions. With regards to treatment options, the respondents showed a positive preference for 8 sessions as compared to 4 sessions. With regards to intervention delivery agents, the respondents had a positive preference for facility nurses as compared to community health volunteers (CHVs). In terms of support, the respondents showed positive preference for parenting skills as compared to peer support. Respondents expressed negative preferences of ANC service combined with older mothers as compared to adolescent friendly services and of being offered refreshments alone. However, a positive preference was revealed for combined refreshments and travel allowance over travel allowance or refreshments alone.

These findings do suggest that young peripartum adolescents would prefer more tailored support that engages them directly but also provides guidance and engagement with their caregivers and partners. There is a strong emphasis on youth friendly maternal and child health care services than being lumped with routine MCH clinics with adult women.

Despite the enormous significance of these findings, these are outputs of a DCE experiment from two Nairobi primary health care sites. This could be tested further in other sites and settings for external validity. In general, DCEs have been shown as effective method for eliciting preferences for mental health services within diverse settings, illustrating a promising approach to increasing patient-centered mental health care.

These directly elicited preferences are consistent with our experience of depression associated challenges that pregnant adolescents experience. We learnt that our respondents preferred longer group psychotherapy and that they did not rank educational needs above vocational training. Pregnancy and impending motherhood may have shaped these preferences which might evolve and it appears that they did see infant care and parenting as their key difficulties. Our respondents would prefer informational support for caregivers and partners above direct involvement of the caregivers and partners in group sessions. It appears that peer support, privacy and a safe space to share their experiences is considered important. Given that a large number of them experience interpersonal disputes with family members and are in conflicted relationship with their partners or partners, something other studies have noted too (45).. It was also clear that they preferred to interact and be serviced in Antenatal clinics that are youth friendly/responsive by nurses but not with older adult women. Pregnant and parenting adolescents differentiate themselves from regular adolescents and adult women [32–34].

A preference of IPT delivery by ANC nurses over CHVs is also telling. While we will now use these revealed preferences as guidance in our efforts to further modify group IPT, we also know that as these young women give birth, their preferences may evolve and change. Keeping a conversation around other aspects of health such as robust SRHR choices-including family planning, use of PrEP, HIV testing, use of contraceptives etc. will also be critical and nurses and CHVs can both play a part there. Working out their livelihood options if return to school is difficult will need to be a priority and for those who will opt for continuation of school, offering brief IPT (4 sessions, even as a booster or remission treatment) might help in the long run. It appears that young pregnant or parenting girls would like to be taken seriously like adult women and would like to access services that are responsive to their needs and offer protection from multidimensional stigma associated with unintended early pregnancy and mental illness.

Depressive disorders are among the top three causes of years lived with disability globally, accounting for 40% of all mental illness, and affecting 350 million people comprising 4% of the population. Mental health services are scarce in low-and-middle-income countries like Kenya. Even when services exist, these do not map on to patient and provider preferences. Innovations are needed to provide accessible, affordable, and acceptable prevention, care, and treatment services to the diverse populations faced with poor mental health. Information and messages about mental health, preventative services, care and treatment characteristics, provider approaches, and care provision modalities must continue to evolve based on stakeholder preferences to ensure relevance and desirability [35].

Historically, patient involvement especially of vulnerable adolescents in shaping health practice has been minimal, especially in low-resource settings (10-13). There is good evidence that services that engage patients from the beginning – around conceptualizing the service itself, can be highly successful and effective. Similarly, adolescent-centered care has been associated with improved symptom burden, satisfaction, and enablement [36].

The current focus on patient-centered care within healthcare systems aims to ensure high-quality interactions between patients and the health system through achievement of the eight principles: *respect for patients’ preferences, coordination and integration of care, information and education, physical comfort, emotional support, involvement of family and friends, continuity and transition, and access to care* [37,38]. It appears that many of these principles were articulated by our respondents in the DCE experiment on depression care.

## Limitations

This study had some limitations. It was conducted in two healthcare facilities in an informal urban setting that are part of Nairobi metropolitan services health facility and results may not be entirely generalizable to other settings and practice models. Therefore, the applications of our findings remain limited to urban informal settlements. These settlements tend to be socioeconomically and ethnically diverse in their own right so the current study can inform more contextual study designs. We studied a convenience sample of respondents who may have been more frequent visitors to the facilities and their views may not represent all patients. Our results and conclusions are based on the attributes and levels included in the DCE we designed. While we followed a robust process to determine which attributes are important and relevant in our context using focus groups of key informants with expert knowledge of the clinical setting as well as previous literature in similar settings, we cannot be sure we captured all important attributes.

## Conclusion

Our participants revealed a complex set of preferences – prioritizing longer psychotherapy duration, parenting support, disseminating relevant depression care information to caregivers and partners as opposed to inviting them into groups, vocational training over return to school and combined refreshments with travel allowance as added incentives for psychotherapy. Negative preferences were revealed for combined ANC services with adult women, and provision of refreshments alone. These directly elicited preferences provide a unique opportunity to develop ‘patient-centered’ mental health services in a primary care context.

## Data Availability

the data underlying the results presented in the study are available from the corresponding author on reasonable request.

## Declarations

### Ethics approval and consent to participate

the study was approved by the Kenyatta National Hospital/University of Nairobi ethical review committee (approval no. P694/09/2018). The study received approval from the Nairobi County health directorate (approval no. CMO/NRB/OPR/VOL1/2019/04) and approval from National Commission for Science, Technology, and Innovation (NACOSTI/P/19/77705/28063). All study participants’ informed consent to participate would be sought, including stakeholders and advisory committee members from whom data would be collected. The research was carried out per the KNH/UoN ethical review committee guidelines as well as the standard guidelines and principles of the Declarations of the Helsinki.

### Consent for publication

All study participants gave their consent to publish this work’s findings.

### Availability of data and material

The datasets generated and/or analyzed during the current study are not publicly available due but are available from the corresponding author on reasonable request.

### Authors’ contributions

MK developed this paper, data collection was overseen by JK, VN, and AT and MQ conducted quantitative data analysis, while OY, SM, KYH, MM, JL, ML, PC and JU.All authors read and approved the work.

## Acknowledgments

The authors would like to thank all the participants, Nairobi County health directorate, Director of Mental health, Ministry of Health, Kariobangi, and Kangemi health facility staff.

**S1-**Supplementary Table 1

**S2-**Supplementary Literature on Attributes and Levels

**S3-**Supplimentary Mixed Logit Models

